# Effectiveness of a Socially Implemented Cognitive Decline Prevention Program: A Retrospective Observational Study

**DOI:** 10.64898/2026.04.08.26350304

**Authors:** Minoru Kouzuki, Keiko Fujita

## Abstract

**Background and Objectives:** Multifactorial interventions have been reported to be effective in improving cognitive function; however, their long-term effectiveness in community settings remains to be sufficiently examined. This study aimed to investigate the effects of a socially implemented multifactorial intervention program on dementia onset, long-term care insurance certification, and post-intervention cognitive and physical functions.

**Methods:** This retrospective observational study collected data from three municipalities. The study population comprised individuals suspected of having mild cognitive decline based on cognitive function screening tests conducted by March 31, 2024, and who had been invited to participate in a dementia prevention class, but had not applied for long-term care insurance at the time of the invitation. Participants were classified into class participation and non-participation groups for analysis. Most participants attended the class only once (intervention duration: 4 or 6 months).

**Results:** Data from 104, 218, and 256 individuals were collected from the three municipalities, respectively. No significant association was found between class participation and suppression of dementia onset or long-term care insurance certification in any of the municipalities. Regarding pre–post comparisons among class participants, significant improvements in cognitive function and some physical functions were observed in all the three municipalities.

**Conclusions:** The multifactorial interventions implemented in community settings showed no effect on dementia onset or health outcomes. However, class participation was associated with improvements in cognitive function and some physical functions. These findings suggest that implementing programs based on evidence can achieve effects similar to those observed in studies conducted under ideal conditions.

## 1. Introduction

Dementia is a multifactorial disorder, and its onset is influenced by numerous modifiable risk factors (Jones et al., 2024; Livingston et al., 2024). To achieve optimal risk reduction, it is important to implement multimodal interventions that target various risk factors, as numerous studies have demonstrated that such approaches improve cognitive function (Castro et al., 2023; Reparaz-Escudero et al., 2024; Rookes et al., 2026; Salzman et al., 2022).

The lead author, in collaboration with relevant stakeholders, developed a program for preventing cognitive decline (named the “Tottori Method Dementia Prevention (TMDP) Program”), which includes physical exercise, cognitive training, and educational lectures on dementia (Kouzuki et al., 2020). This program was implemented as a 6-month intervention comprising weekly 2-h sessions (50 min of physical exercise, 20 min of lectures or breaks, and 50 min of cognitive training) targeting community-dwelling individuals aged ≥65 years with suspected mild cognitive decline. Consequently, improvements in cognitive and some physical functions were reported (Kouzuki et al., 2020). This initiative, started with the aim of disseminating the program to regions without researchers, was evaluated using a framework in which participant selection, intervention delivery, and primary evaluation can be conducted by municipalities. The contents of physical exercise and lectures are recorded on DVDs, and pamphlets are prepared for cognitive training; these materials are provided free of charge upon request to facilitate use in various settings (*‘Tottori Prefecture Web site’*, n.d.). As a result of efforts to disseminate these research findings since fiscal year 2019, initiatives for dementia prevention using this program have been implemented in various regions in Tottori Prefecture, Japan.

The goal of engaging in dementia prevention activities is to avoid developing dementia. Multifactorial lifestyle interventions considered effective for preventing cognitive decline prevent or reduce frailty and maintain activities of daily living (Kulmala et al., 2019; Pöyhönen et al., 2025; Saarela et al., 2025). These results suggest long-term benefits in reducing disability and the need for long-term care; however, sufficient evidence demonstrating the efficacy of multifactorial interventions in reducing dementia incidence is lacking (Hafdi et al., 2021). In particular, the effectiveness of the TMDP Program in reducing the risk of dementia onset and other long-term health outcomes remains to be examined. Furthermore, even if an intervention has demonstrated efficacy under ideal research conditions, various barriers hinder its implementation in real-world settings (Lau et al., 2016), leading to a gap between research evidence and practice. Therefore, the effectiveness of the program should be evaluated in community settings without direct involvement of researchers.

In this study, we obtained data from municipalities that implemented dementia prevention classes using the TMDP Program and examined whether participation in the classes was associated with differences in dementia incidence and long-term care insurance (LTCI) certification among individuals with mild cognitive decline. In addition, changes in cognitive and physical functions were examined.

## 2. Materials and methods

### 2.1. Study design

This retrospective observational study was conducted in Misasa, Hokuei, and Yurihama Towns and data were collected between July 2024 and March 2025. The study was approved by the Ethical review board, Faculty of Medicine Tottori University (No. 24A040; June 13, 2024). As this was a non-invasive, non-interventional observational study using retrospective data, the need for consent was waived by the Ethical review board. Instead, an opt-out method was employed to ensure that potential subjects had the opportunity to refuse participation. Information regarding the study was disclosed via the website of the Tottori University Hospital. For data collection involving residents of Misasa Town, notifications were published on the municipal bulletin board and sent directly to the subjects; for residents of Hokuei and Yurihama Towns, the information was disclosed on the respective official websites of the municipal offices. All procedures were conducted with strict respect for the privacy and confidentiality of the participants.

### 2.2 Study participants

The study fields comprised three Towns (Misaas, Hokuei, and Yurihama) operating dementia prevention classes as public services based on the subject selection methods and programs of previous research (Kouzuki et al., 2020). Participants were individuals suspected of mild cognitive decline via the Monowasure Soudan Program (MSP) (Inoue et al., 2009) —a brief cognitive screening test—conducted by municipal offices (Misasa Town office and Hokuei Town office: April 1, 2019, to March 31, 2024; Yurihama Town Hall: April 1, 2020, to March 31, 2024), who received invitations to the dementia prevention class, and who had not applied for LTCI certification (Supplementary Table 1). In all towns, individuals who declined the use of their data were excluded from the study.

### 2.3 Data collection

In the three Towns, classes were conducted as shown in Table 1. Although some individuals attended multiple classes in each municipality, participants were newly recruited for each class; therefore, most individuals participated in only one class (intervention duration: 4 or 6 months).

**Table 1.**
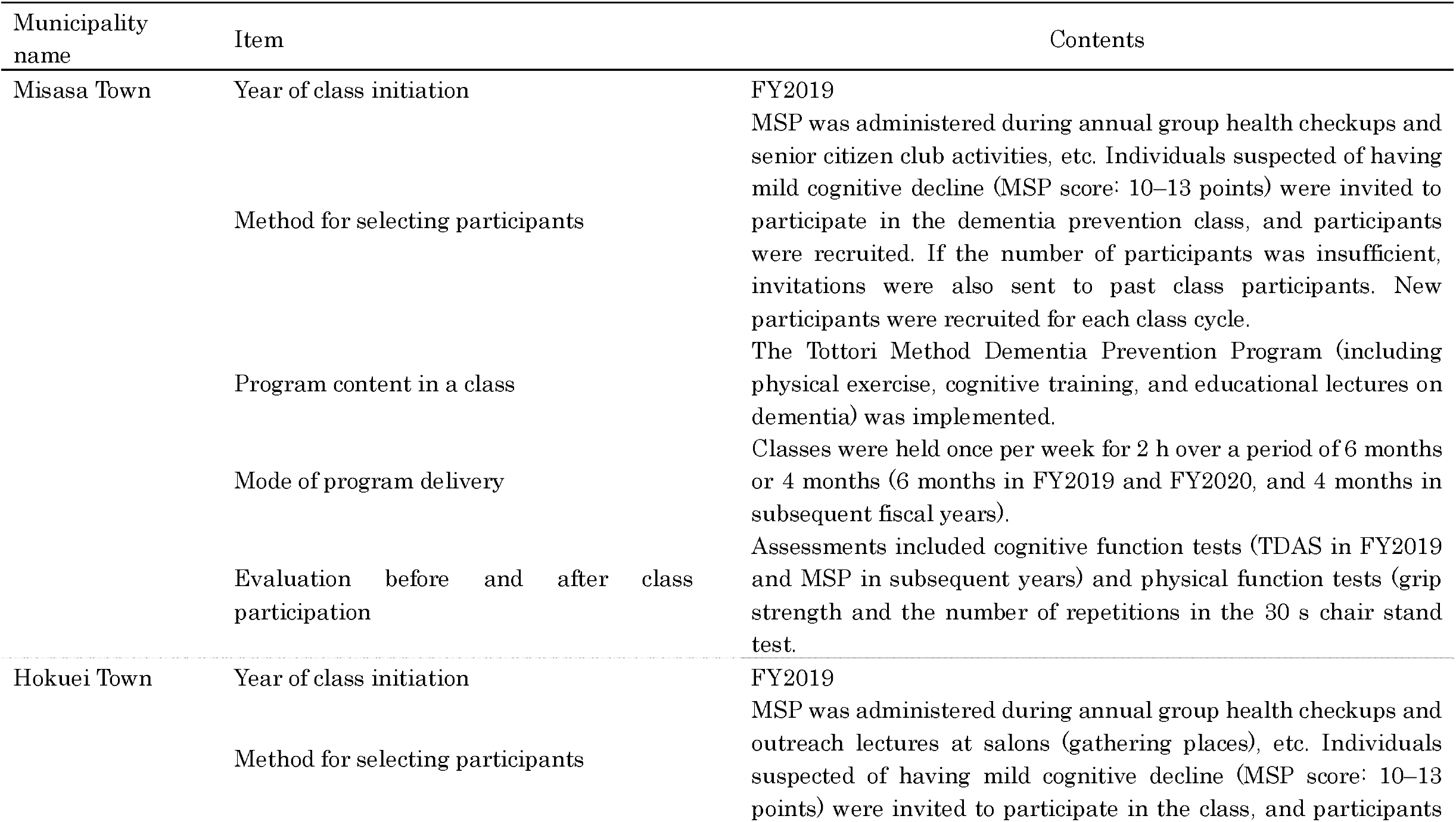
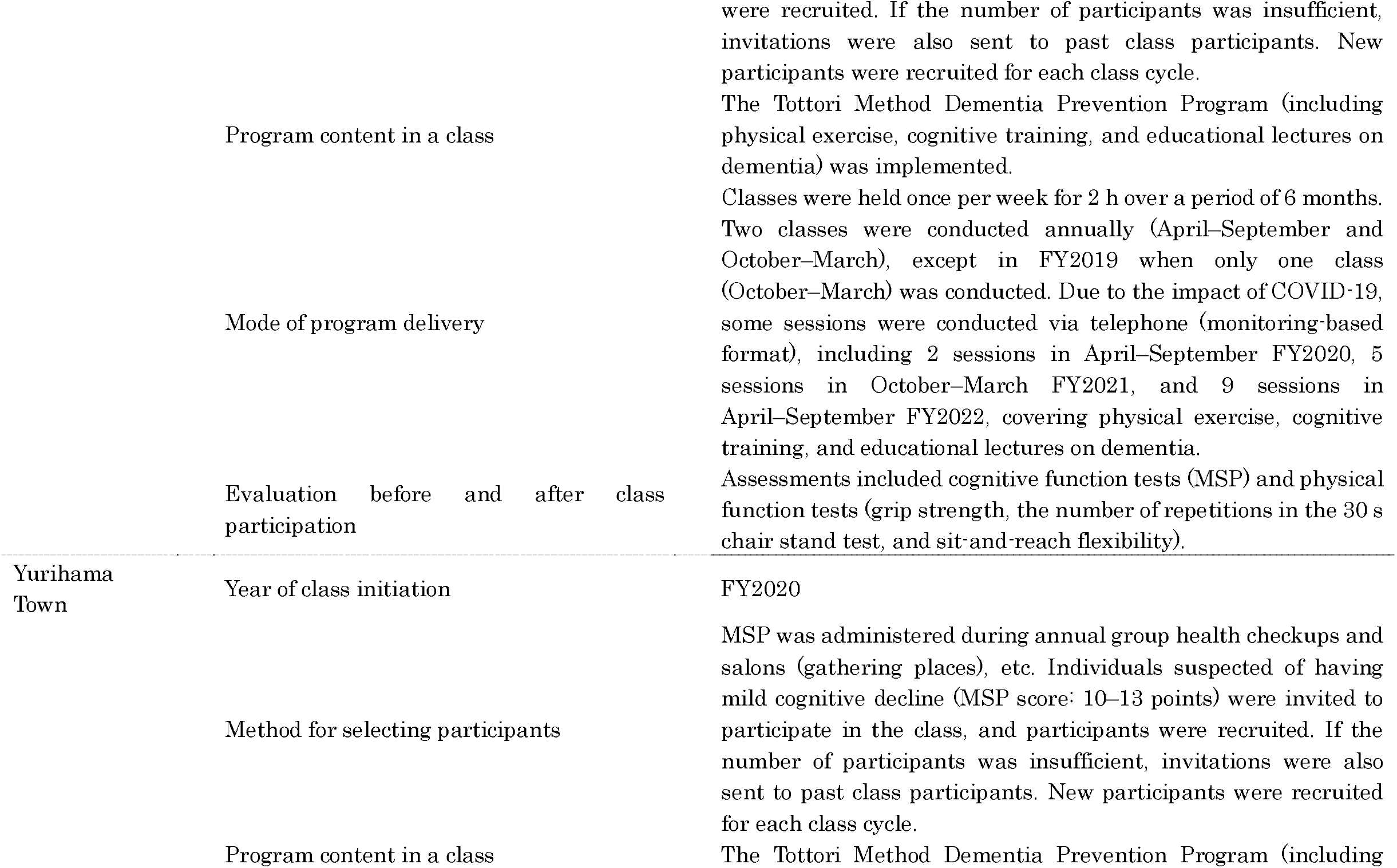
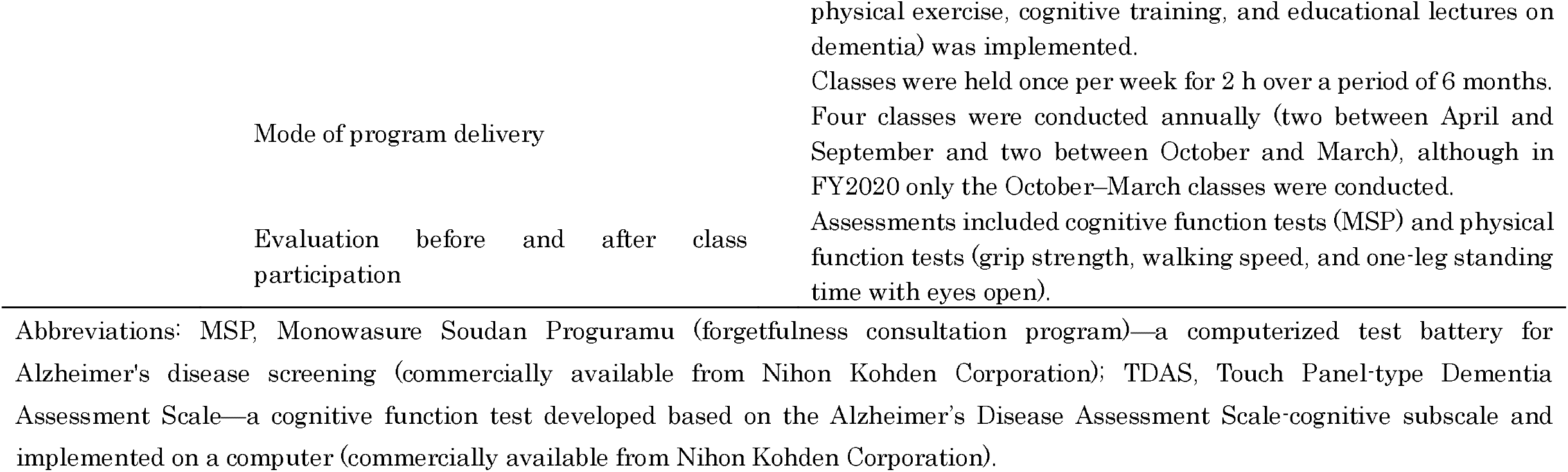
Operational methods of dementia prevention class in each municipality.

The data included participant background characteristics (age and sex), participation in the dementia prevention class (including year of participation and number of times attended per class), LTCI–related information (presence or absence of certification, date of certification, and whether the diagnosis described in the physician’s opinion document was dementia), survival (date of death for deceased individuals), relocation outside the town, cognitive function assessment results, and physical function assessment results.

For cognitive function assessments, data on the MSP (Nihon Kohden Corporation, Tokyo, Japan) (Inoue et al., 2009) and Touch Panel-type Dementia Assessment Scale (TDAS) (Nihon Kohden Corporation, Tokyo, Japan) (Inoue et al., 2011), including dates and results, were collected. The MSP is a simple cognitive screening test deployed on a touch-panel computer and comprises four items. A total score (0–15 points) is calculated, with lower scores indicating more severe cognitive impairment. In addition to MSP results obtained during screening for cognitive impairment in each municipality, MSP results measured before and after class participation were collected for participants in the dementia prevention classes. If cognitive function testing was not conducted before class participation, the screening test results used for participant selection were regarded as the pre-intervention values. The TDAS is a cognitive function test developed based on the Alzheimer’s Disease Assessment Scale–cognitive subscale (Rosen et al., 1984) and implemented on a touch-panel computer. It comprises nine items, and a total score (0–101 points) is calculated, with higher scores indicating more severe cognitive impairment. TDAS results were collected before and after class participation. However, this test was not conducted in Hokuei and Yurihama Towns.

Physical function assessment data, including the dates and results of assessments conducted before and after class participation, were collected for individuals who participated in the dementia prevention classes. The collected items included grip strength (the larger value between the left and right hands), number of repetitions in the 30 s chair stand test, the sit-and-reach score, one-leg standing time with eyes open (the larger value between the left and right sides, up to 60 s), and walking speed (5-m walking time). However, the measurement items differed by municipality: grip strength and number of repetitions in the 30 s chair stand test were collected in Misasa Town; grip strength, number of repetitions in the 30 s chair stand test, and sit-and-reach score were collected in Hokuei Town; and grip strength, walking speed, and one-leg standing time with eyes open were collected in Yurihama Town.

### 2.4 Statistical analysis

Statistical analyses were performed using SPSS version 27 (IBM Corporation, Tokyo, Japan) and EZR version 1.55 (Saitama Medical Center, Jichi Medical University, Saitama, Japan) (Kanda, 2013). Statistical significance was set at p < 0.05. Analyses were conducted using complete cases for each relevant variable, that is, only available data without missing values were used for each analysis.

Individuals who attended the classes were defined as those who participated in the class for at least one day, whereas individuals who did not attend the classes were defined as those who underwent MSP between April 1, 2019, and March 31, 2024, and never participated in the classes thereafter. Because including individuals with less participation in the participation group was considered inappropriate, additional analyses were conducted excluding individuals with low attendance (defined as attendance of one-third or less of the scheduled sessions) and those who discontinued participation (defined as absence for three or more consecutive sessions with no subsequent return). However, data on those who discontinued participation were available only for Misasa Town.

Baseline characteristics were compared between the two groups using the following methods: (1) for continuous variables, normality was assessed using the Shapiro–Wilk test, and after confirming the equality of variance using Levene’s test, Student’s t-test or the Mann–Whitney U test was used; (2) for categorical variables, Fisher’s exact test was used.

The association between participation in dementia prevention classes and dementia onset or LTCI certification was analyzed as follows. In all analyses, for individuals who attended the classes, the start date of follow-up was defined as the date of assessment of cognitive or physical function conducted before class participation (if these assessments were conducted on different dates, the later date was used; for individuals who attended multiple classes, the date of the pre-participation assessment for the first class was used). For individuals who did not attend the classes, the start date of follow-up was defined as the date of the first MSP assessment. The end of follow-up was March 31, 2024. Dementia onset was determined based on information in the physician’s opinion document submitted for LTCI application.

Incidence rates (100 person-years) of dementia onset and LTCI certification were estimated, and Kaplan–Meier analyses were performed. Survival curves were compared between the participation and non-participation groups using the log-rank test. In addition, Cox proportional hazards models were used to evaluate the association between class participation and dementia onset or LTCI certification. Dementia onset and LTCI certification were treated as dependent variables and class participation as the independent variable. Hazard ratio (HR) and 95% confidence interval (95% CI) were calculated (the calculation of 95% CI was performed as a post hoc analysis not specified in the original study protocol). Because the number of outcome events per independent variable was small, analyses adjusted for covariates were not performed, as the results from the fitted proportional hazards regression model may not be valid (Peduzzi et al., 1995).

To compare cognitive and physical function before and after participation in the dementia prevention classes, normality was assessed using the Shapiro–Wilk test, and paired t-tests or Wilcoxon signed-rank tests were used as appropriate. Because some individuals participated in multiple classes, multiple data points from the same individual may have been included. Therefore, analyses were conducted using both per-person data (only one dataset per individual) and per-participation data (using multiple data points even for a single individual), and robustness was assessed. For per-person analyses, data from the first class attended were used. For per-participation analyses, if the MSP score at the time of participation fell outside the range of 10–13 during repeated participation, the data from those sessions were excluded from the analysis.

## 3. Results

### 3.1 Results for Misasa Town

Data from 104 individuals, excluding two individuals who met the exclusion criteria, were collected and analyzed. Among them, 24 were class participants, and 80 were non-participants. Compared with non-participants, participants had a higher proportion of women (p = 0.030) (Table 2). No significant association was observed between class participation and either dementia onset or LTCI certification (Supplementary Figure 1, Table 3). Regarding changes in cognitive and physical functions before and after class participation (Figure 1, Supplementary Tables 2–4), analyses were conducted separately for participants in fiscal year 2019, fiscal year 2020, and fiscal years 2021–2023, because the intervention period and evaluation items differed over time. As a result, in the analysis using cumulative per-participation data for fiscal years 2021–2023, significant improvements were observed in MSP scores and number of repetitions in the 30 s chair stand test (p = 0.035 and p = 0.016, respectively).

**Table 2.**
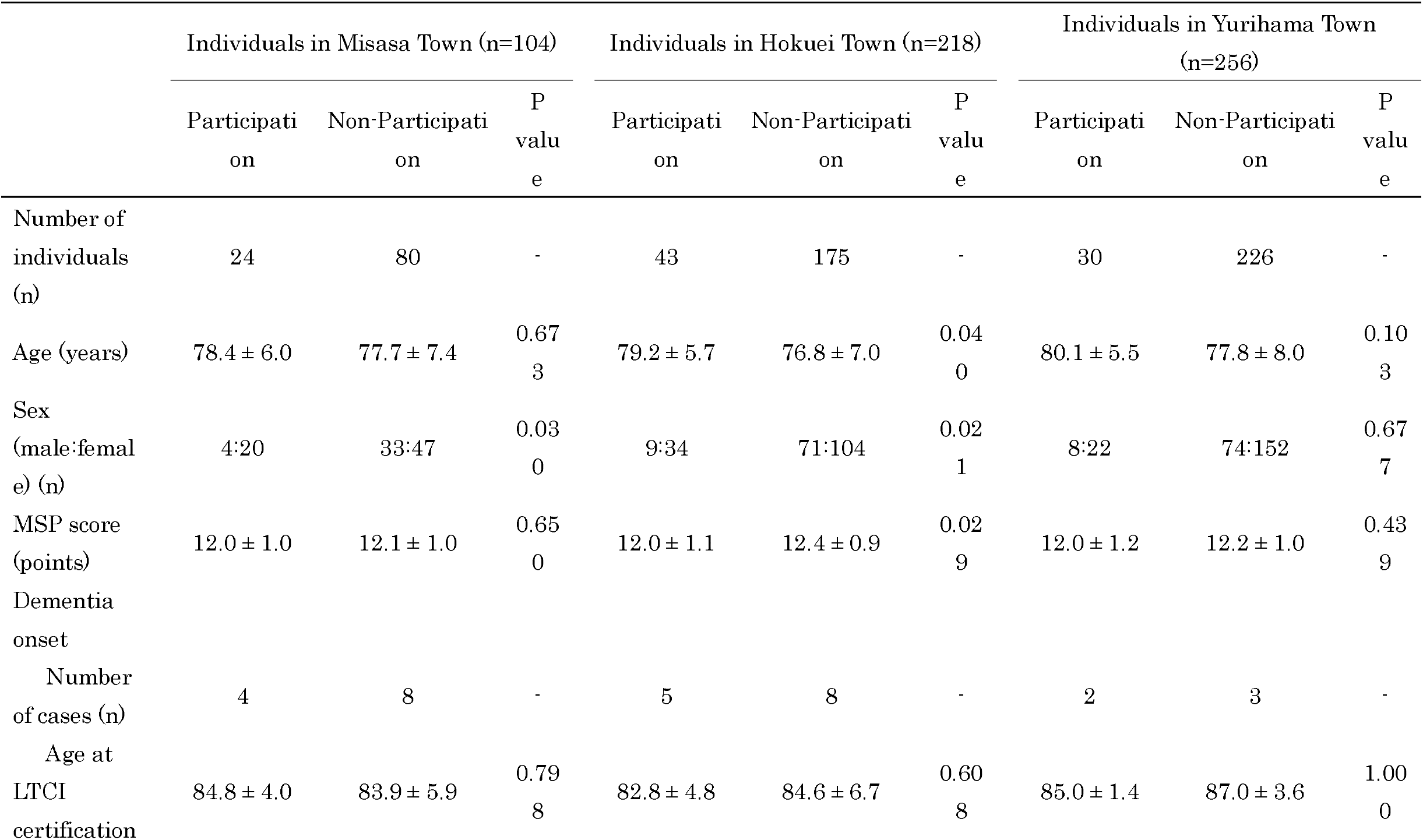
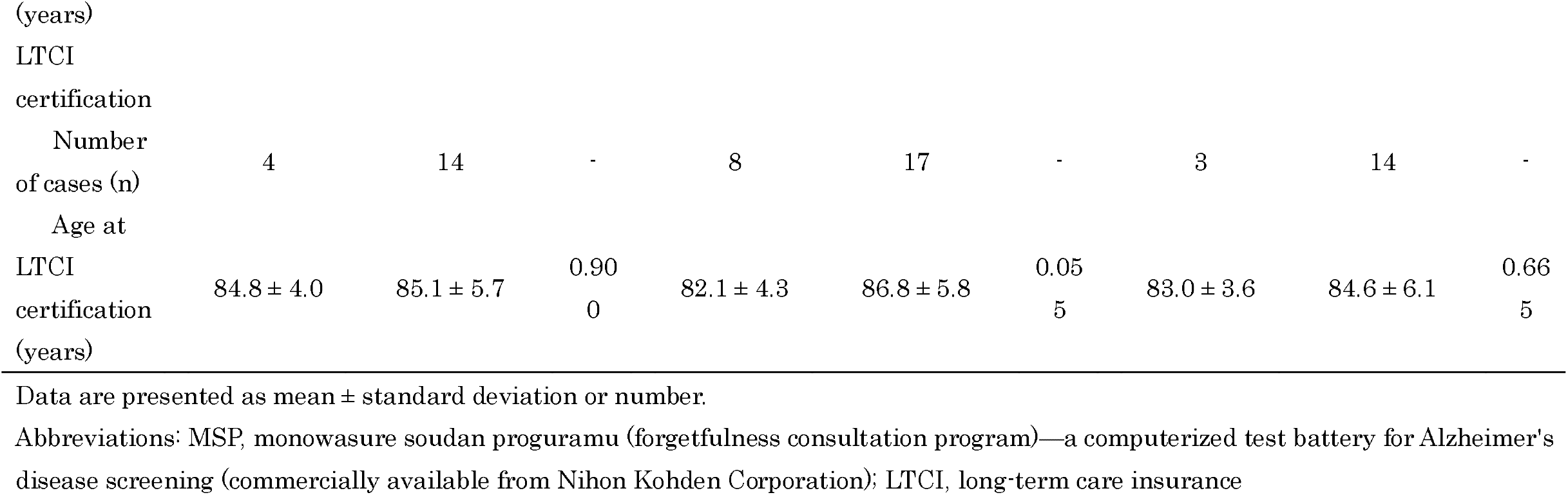
Characteristics of target populations in each municipality.

**Table 3.**
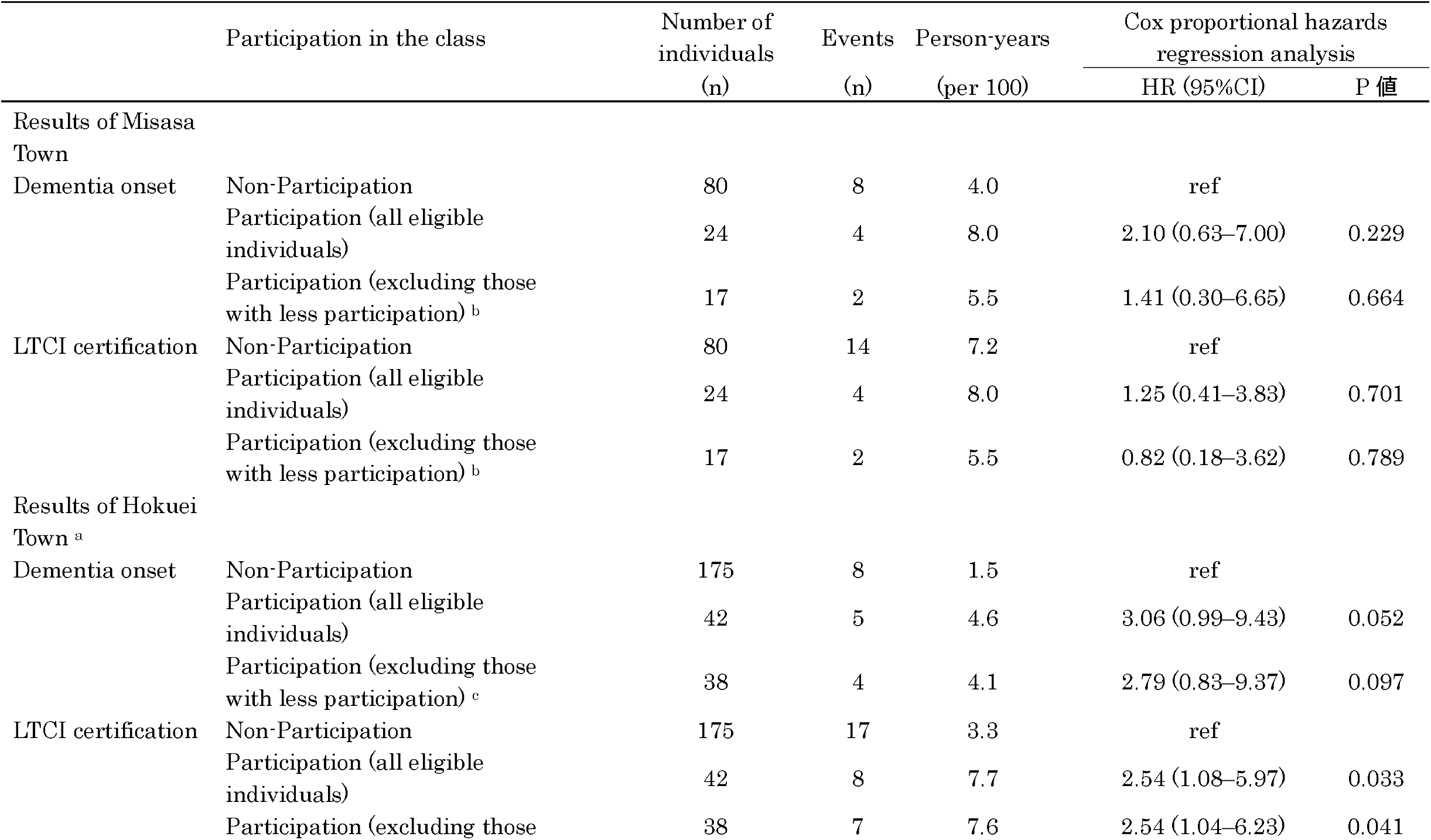
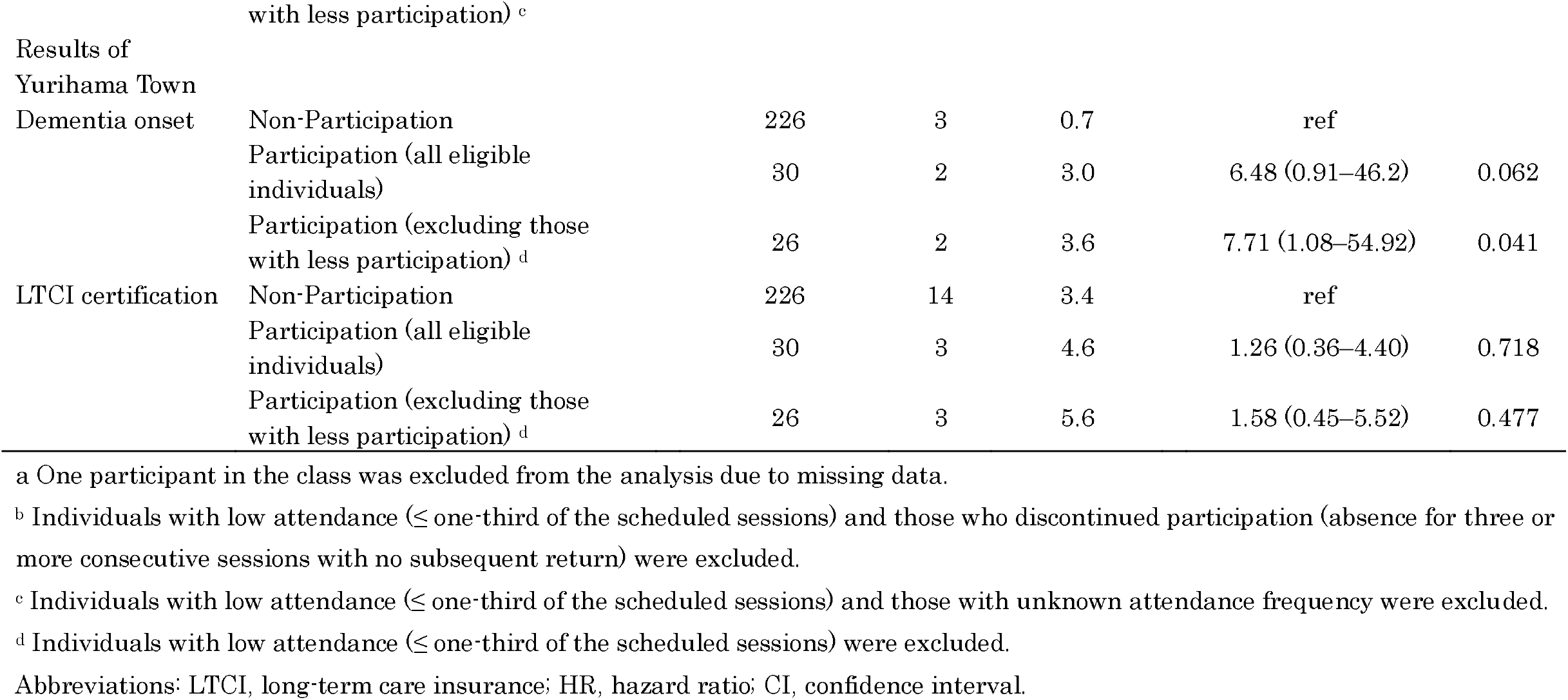
Impact of class participation on the onset of dementia and eligibility for long-term care insurance.

**Figure 1.**
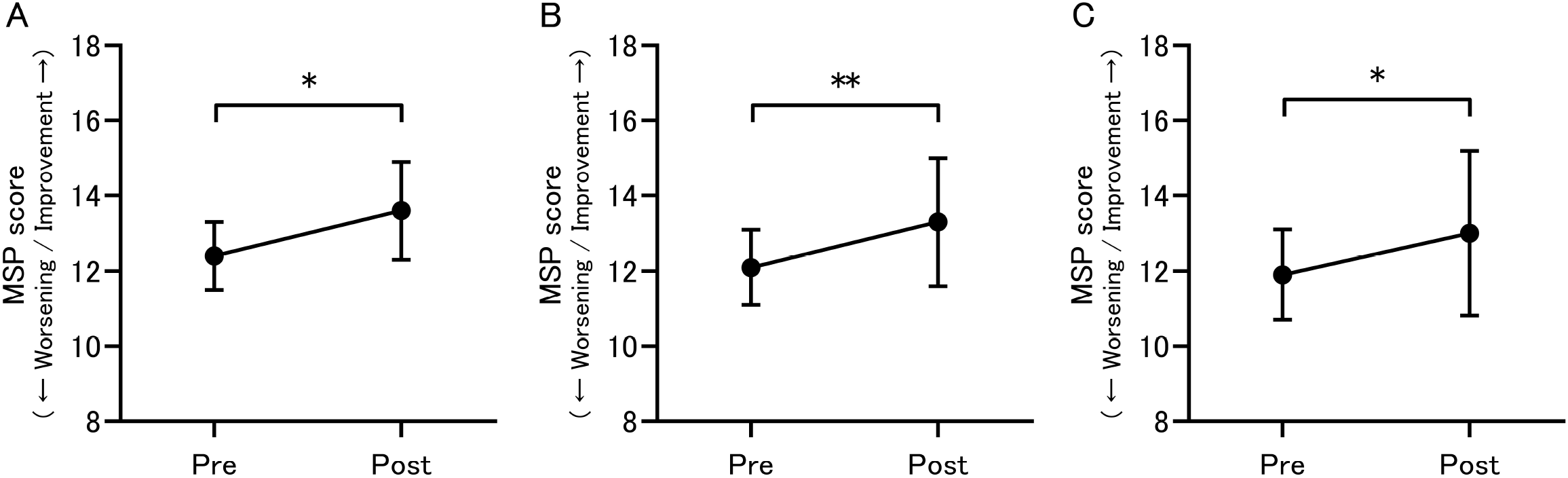
Changes in cognitive function before (pre) and after (post) class participation based on an analysis using cumulative per-participation data (including multiple records per individual). (A) Participants in Misasa Town (n = 14), (B) participants in Hokuei Town (n = 42), and (C) participants in Yurihama Town (n = 29). Data are expressed as mean ± standard deviation. * p<0.05, ** p<0.01 Abbreviations: MSP, Monowasure Soudan Proguramu (forgetfulness consultation program)—a computerized test battery for Alzheimer’s disease screening (commercially available from Nihon Kohden Corporation)

### 3.2 Results for Hokuei Town

In total, 218 individuals were recruited, among whom 43 were class participants and 175 were non-participants. Compared with non-participants, participants were older (p = 0.040), had a higher proportion of women (p = 0.021), and had lower cognitive function test scores (p = 0.029) (Table 2). Class participation was significantly associated with LTCI certification in both the analysis including all participants and that excluding participants with low attendance (HR = 2.54, 95% CI = 1.08–5.97; HR = 2.54, 95% CI = 1.04–6.23, respectively) (Supplementary Figure 2, Table 3). Regarding changes in cognitive and physical functions before and after class participation (Figure 1, Supplementary Table 5), analysis based on cumulative per-participation data showed that MSP scores significantly improved (p < 0.001), whereas grip strength significantly decreased (p = 0.001). In the analysis excluding individuals with low attendance, significant improvements were observed in MSP scores and number of repetitions in the 30 s chair stand test (p < 0.001 and p = 0.026, respectively), whereas grip strength showed a significant decrease (p = 0.002).

### 3.3 Results for Yurihama Town

In total, 256 individuals were recruited, among whom 30 were class participants and 226 were non-participants. No significant differences in baseline characteristics were observed between the participants and non-participants (Table 2). In the analysis excluding individuals with low attendance, class participation was significantly associated with dementia onset (HR = 7.71, 95% CI = 1.08–54.92) (Supplementary Figure 3, Table 3). Regarding changes in cognitive and physical functions before and after class participation (Figure 1, Supplementary Table 6), analysis based on cumulative per-participation data showed significant improvement in MSP scores (p = 0.031).

## 4. Discussion

In this study, we evaluated the effectiveness of a cognitive decline prevention program that had previously demonstrated efficacy under research conditions. In a previous study (Kouzuki et al., 2020), improvements in cognitive and some physical functions were observed after a 6-month intervention; however, in the present study, no effects on reducing the incidence of dementia or LTCI certification were observed. Nevertheless, class participation was associated with improvements in cognitive and some physical functions, indicating that evidence-based implementation can achieve effects similar to those observed in studies conducted under ideal conditions. Furthermore, this study comprised independent analyses in three municipalities, and the consistency of the results supports the robustness and generalizability of the findings. To the best of our knowledge, this study is among the first to demonstrate the social implementation of initiatives, encompassing appropriate participant selection, intervention delivery, and evaluation, to prevent cognitive decline.

Across the three municipalities, participation in classes providing the Tottori Method Dementia Prevention Program for individuals with mild cognitive decline did not result in lower rates of dementia onset or LTCI certification compared with non-participation. Therefore, this analysis did not identify long-term beneficial effects of short-term class participation. Similarly, previous studies examining the long-term effects of interventions on dementia incidence have also not demonstrated significant reductions in dementia incidence (Coley et al., 2025; Moll van Charante et al., 2016; Sink et al., 2015). Moreover, although short-term interventions can delay the progression of disability, their effects tend to diminish over time after the intervention period ends (Park et al., 2021; Szanton et al., 2019). The results of the present study are consistent with these previous reports. Several factors may explain the absence of long-term effects in this study. First, dementia onset was determined based on information from the physicians’ opinion documents submitted for LTCI applications; however, some individuals may not apply for LTCI certification after dementia diagnosis, and thus not all cases of dementia onset were captured. Similarly, for health conditions other than dementia, because the LTCI certification was used as the outcome, individuals with health impairments who did not apply for the LTCI were not captured. Second, the incidence of dementia in the non-participation group was relatively low. For instance, the incidence of dementia in this study was lower than that in studies targeting individuals with mild cognitive impairment (Ding et al., 2016; Ishikawa & Ikeda, 2007; Solfrizzi et al., 2004; Zhang et al., 2021). Individuals who undergo cognitive function screening may have a higher level of interest in dementia and its prevention, and undergoing screening may lead to behavioral changes. Indeed, health information acquisition, understanding, and application significantly influence health awareness among older adults, thus impacting their health outcomes (Li et al., 2025). Furthermore, contrary to expectations, some municipalities had a higher rate of care certifications due to dementia or other reasons among the class participants. This finding may reflect earlier utilization of long-term care services facilitated by connections with municipal support systems. Social participation is reported to alleviate the risk of dementia and improve population health (Fujihara et al., 2025; Ide et al., 2026; Taniguchi & Ukawa, 2022). One possible mechanism is that social connections enable individuals to receive various forms of support (Steijvers et al., 2023; Umberson et al., 2010). Thus, participation in classes may have led to increased access to information and connections with municipal staff, resulting in earlier applications for LTCI certification. Appropriate utilization of long-term care services is beneficial for improving quality of life (Wilby et al., 2013). Therefore, to draw more accurate conclusions, further investigation of care level classifications is warranted.

Among class participants, significant improvements in cognitive function and some physical functions were observed. These findings suggest that implementing programs based on scientific evidence in realistic settings can achieve effects comparable to those observed in research environments. Regarding physical function, although many lower-extremity-related measures showed improvements, no significant improvement in grip strength was observed. This finding differs from a previous report that showed improvement in grip strength (Kouzuki et al., 2020). The reason for this finding remains unclear. However, a meta-analysis examining the effectiveness of a multicomponent exercise in older adults with cognitive frailty showed more significant improvements in lower limb strength as opposed to grip strength, which the authors attributed to multicomponent exercise protocols emphasizing lower limb muscle training while leaving upper limb muscle exercises underrepresented (Luo et al., 2024). Given that strength-focused hand training results in meaningful improvements in grip strength (Akbaş, 2025), the TMDP Program may benefit from incorporating components targeting upper extremity strength.

This study has several limitations. First, dementia onset and health impairments could not be accurately determined for all participants. Second, the intervention period was relatively short (4–6 months), and this study did not evaluate the effects of long-term continuous interventions over several years. Given that exercise detraining may negatively affect cognitive function (Ferreira et al., 2024), short-term interventions may be insufficient to achieve long-term effects. Strategies are needed to encourage continued engagement in preventive activities after the program ends within the constraints of limited social resources, such as budget and personnel. Third, some participants had relatively short follow-up periods. Although the maximum follow-up period was 5 years, some participants were followed for less than 1 year, which may have resulted in a low number of dementia onset and LTCI certification events. In addition, although many factors are known to be associated with dementia onset and functional disability (Cheng et al., 2025; Jones et al., 2024; Livingston et al., 2024; Stuck et al., 1999), the available data in this study were limited, and sufficient adjustment for these factors was not possible. Moreover, because the number of events was small, analyses adjusted for covariates were not performed; however, as the mean age of participants was higher than that of non-participants, the effect of age on the results cannot be ruled out.

In conclusion, short-term participation in a scientifically validated dementia prevention program implemented in community settings showed no effect on long-term dementia incidence or health outcomes. However, by selecting participants and delivering the program based on previous research, improvements in cognitive and some physical functions, comparable to those observed in research settings, were achieved. These findings highlight the importance of optimizing the implementation process of community-based cognitive decline prevention interventions. Future studies should focus on establishing support systems that enable continued engagement in preventive activities after completion of the dementia prevention class. In addition, studies with longer intervention and follow-up periods should be conducted to evaluate the preventive effects on dementia and health outcomes.

## Data Availability

The data sets used and/or analyzed during the current study are available from the corresponding author on request. However, access to the data sets will only be granted upon approval by the data provider (the Welfare Division, Misasa Town; Social Welfare Division, Hokuei Town Office; and Welfare Division, Yurihama Town) and the ethics committee.

## CRediT authorship contribution statement

Minoru Kouzuki: Conceptualization, Data curation, Formal analysis, Funding acquisition, Methodology, Project administration, Visualization, Writing – original draft, Writing – review and editing. Keiko Fujita: Investigation, Data curation, Writing – review and editing.

## Acknowledgments

We would like to thank the staff of the Welfare Division, Misasa Town; Social Welfare Division, Hokuei Town Office; and Welfare Division, Yurihama Town for their cooperation during this study. We also thank Editage (www.editage.com) for the English language editing.

## Funding

This work was supported by JSPS KAKENHI [Grant No. JP22K17371]. The funding body was not involved in the study design; collection, analysis, and interpretation of data; writing of the report; and decision to submit the article for publication.

## Declaration of competing interest

The authors declare no conflict of interest. One of the co-authors is affiliated with the municipality that provided the data for this study; however, this author was not involved in the data analysis.

## Declaration of generative AI and AI-assisted technologies in the writing process

ChatGPT (OpenAI), Gemini (Google), and Claude (Anthropic) were used to assist in drafting and language editing of this manuscript. The author critically reviewed and revised the output and take full responsibility for the final content.

## Notes

### Author Declarations

The study was approved by the Ethical review board, Faculty of Medicine Tottori University (No. 24A040; June 13, 2024).

