## Supplemental Material for "Effectiveness of a Socially Implemented Cognitive Decline Prevention Program: A Retrospective Observational Study"

Table S1. Detailed inclusion criteria for participants in each municipality

| | Inclusion criteria |
|---|---|
| Individuals for whom data were provided by Misasa Town Office | 1) Individuals suspected of having mild cognitive decline based on a simple cognitive function screening test using a touch-panel computer (MSP score: 10–13 points), conducted by the Misasa Town Office during group health checkups, at senior citizen clubs and salons, and during home visits between April 1, 2019, and March 31, 2024.<br>2) Individuals who received an invitation to participate in a dementia prevention class conducted by the Misasa Town Office (among those with MSP scores of 10–13, information about the classes were not provided to those already using long-term care services, those judged by the municipal office to be extremely elderly, or those without means to attend the venue).<br>3) Individuals who had not applied for long-term care insurance at the time they first received an invitation to participate in the dementia prevention class.<br>※Individuals who underwent the MSP multiple times within the same fiscal year and had an MSP score ≥14 before attending the class in that fiscal year were excluded, as they were not considered to have mild cognitive decline. Such cases would involve initiating follow-up from a cognitively normal stage and were therefore deemed inappropriate for this study, which aimed to evaluate outcomes among individuals with cognitive decline. |
| Individuals for whom data were provided by Hokuei Town Office | 1) Individuals suspected of having mild cognitive decline based on a simple cognitive function screening test using a touch-panel computer (MSP score: 10–13 points), conducted by the Hokuei Town Office during group health checkups, salon outreach lectures, and during home visits between April 1, 2019, and March 31, 2024.<br>2) Individuals who received an invitation to participate in a dementia prevention class conducted by the Hokuei Town Office (among those with MSP scores of 10–13, information about the classes were not provided to those already participating in programs for long-term care [functional disabilities] prevention or using long-term care services).<br>3) Individuals who had not applied for long-term care insurance at the time they first received an invitation to participate in the dementia prevention class.<br>※Individuals who underwent the MSP multiple times and had an MSP score ≥14 before attending the class were |

| | |
|---|---|
| | excluded, as they were not considered to have mild cognitive decline. Such cases would involve initiating follow-up from a cognitively normal stage and were therefore deemed inappropriate for this study, which aimed to evaluate outcomes among individuals with cognitive decline. |
| Individuals for whom data were provided by Yurihama Town Hall | 1) Individuals suspected of having mild cognitive decline based on a simple cognitive function screening test using a touch-panel computer (MSP score: 10–13 points) conducted by the Yurihama Town Hall during group health checkups, at salons, and during home visits between April 1, 2020, and March 31, 2024.<br>2) Individuals who received an invitation to participate in a dementia prevention class conducted by the Yurihama Town Hall (among those with MSP scores of 10–13, information about the classes were not provided to those already using long-term care services).<br>3) Individuals who had not applied for long-term care insurance at the time they first received an invitation to participate in the dementia prevention class.<br>※Individuals who underwent the MSP multiple times and had an MSP score ≥14 before attending the class were excluded, as they were not considered to have mild cognitive decline. Such cases would involve initiating follow-up from a cognitively normal stage and were therefore deemed inappropriate for this study, which aimed to evaluate outcomes among individuals with cognitive decline. |

Abbreviations: MSP, monowasure soudan proguramu (forgetfulness consultation program)—a computerized test battery for Alzheimer's disease screening (commercially available from Nihon Kohden Corporation)

Table S2. Participant characteristics and test results in the comparison of pre- and post-intervention assessments (Misasa Town, fiscal year 2019)

| | Analysis of unique individuals | | |
|---|---|---|---|
| | Pre | Post | P value |
| Number of individuals (n) | 2 | | - |
| Age (years) | 84.5 ± 2.1 | | - |
| Sex (male:female) (n) | 1:1 | | - |
| Class participation (24 sessions) | | | |
| Frequency (times) | 14.0 ± 2.8 | | - |
| Percentage (%) | 58.3 ± 11.8 | | - |
| TDAS score (points) | 11.5 ± 9.2 | 8.0 ± 0 | 0.655 |

Data are presented as mean ± standard deviation or number.

Note: No individuals had low attendance (≤ one-third of the scheduled sessions) or discontinued participation (absence for three or more consecutive sessions with no subsequent return).

Abbreviations: TDAS, Touch Panel-type Dementia Assessment Scale—a cognitive function test developed based on the Alzheimer's Disease Assessment Scale-cognitive subscale and implemented on a computer (commercially available from Nihon Kohden Corporation).

Table S3. Participant characteristics and test results in the comparison of pre- and post-intervention assessments (Misasa Town, fiscal year 2020)

| | Analysis of unique individuals | | |
|---|---|---|---|
| | Pre | Post | P value |
| Number of individuals (n) | 5 | | - |
| Age (years) | 77.0 ± 5.9 | | - |
| Sex (male:female) (n) | 1:4 | | - |
| Class participation (Total 24 sessions) | | | |
| Frequency (times) | 18.6 ± 2.4 | | - |
| Percentage (%) | 77.5 ± 10.0 | | - |
| MSP score (points) | 12.2 ± 1.1 | 13.8 ± 0.8 | 0.066 |
| Grip strength (kg) | 27.4 ± 6.6 | 28.7 ± 7.2 | 0.343 |
| Male (kg) | 39.0 | 40.0 | - |
| Female (kg) | 24.5 ± 1.2 | 25.9 ± 4.0 | 0.472 |
| Sit-to-stand test (n/30 s) | 16.4 ± 6.4 | 20.0 ± 4.5 | 0.062 |

Data are presented as mean ± standard deviation or number.

Note: No individuals had low attendance (≤ one-third of the scheduled sessions) or discontinued participation (absence for three or more consecutive sessions with no subsequent return).

Abbreviations: MSP, monowasure soudan proguramu (forgetfulness consultation program)—a computerized test battery for Alzheimer's disease screening (commercially available from Nihon Kohden Corporation)

Table S4. Participant characteristics and physical function test results in the comparison of pre- and post-intervention assessments (Misasa Town, fiscal years 2021–2023)

| | Analysis of unique individuals | | | Analysis of total observations | | |
|---|---|---|---|---|---|---|
| | Pre | Post | P value | Pre | Post | P value |
| Number of individuals (n) | 13 | | - | 14 | | - |
| Age (year) | 78.5 ± 6.3 | | - | 78.7 ± 6.1 | | - |
| Sex (male:female) (n) | 1:12 | | - | 1:13 | | - |
| Class participation (Total 16 sessions) | | | | | | |
| Frequency (times) | 12.0 ± 2.5 | | - | 12.1 ± 2.5 | | - |
| Percentage (%) | 75.0 ± 15.7 | | - | 75.9 ± 15.5 | | - |
| MSP score (points) | 12.4 ± 0.9 | 13.8 ± 1.3 | 0.033 | 12.4 ± 0.9 | 13.6 ± 1.3 | 0.035 |
| Grip strength (kg) | 24.6 ± 4.6 | 23.4 ± 3.6 | 0.030 | 24.5 ± 4.5 | 23.6 ± 3.6 | 0.117 |
| Male (kg) | 33.9 | 31.1 | - | 33.9 | 31.1 | - |
| Female (kg) | 23.8 ± 3.8 | 22.7 ± 2.9 | 0.060 | 23.7 ± 3.7 | 23.0 ± 3.0 | 0.202 |
| Sit-to-stand test (n/30 s) | 20.2 ± 5.8 | 23.6 ± 8.5 | 0.015 | 20.1 ± 5.6 | 23.3 ± 8.3 | 0.016 |

Data are presented as mean ± standard deviation or number.

Note: The analysis was conducted using only data from FY2021 onward, for which the assessment items before and after class participation and the class duration were consistent.

No individuals had low attendance (≤ one-third of the scheduled sessions) or discontinued participation (absence for three or more consecutive sessions with no subsequent return).

Abbreviations: MSP, monowasure soudan proguramu (forgetfulness consultation program)—a computerized test battery for Alzheimer's disease screening (commercially available from Nihon Kohden Corporation)

Table S5. Participant characteristics and physical function test results in the comparison of pre- and post-intervention assessments (Hokuei Town)

| | Analysis of unique individuals [a] | | | Analysis of total observations | | | Analysis of total observations + Analysis excluding individuals with less participation | | |
|---|---|---|---|---|---|---|---|---|---|
| | Pre | Post | P value | Pre | Post | P value | Pre | Post | P value |
| Number of individuals (n) | 36 | | - | 42 | | - | 41 | | - |
| Age (years) | 79.0 ± 5.3 | | - | 79.4 ± 5.5 | | - | 79.3 ± 5.5 | | - |
| Sex (male:female) (n) | 7:29 | | - | 9:33 | | - | 9:32 | | - |
| Class participation (Total 24 sessions) | | | | | | | | | |
| Frequency (times) | 19.7 ± 4.3 | | - | 19.9 ± 4.2 | | - | 19.9 ± 4.2 | | - |
| Percentage (%) | 82.2 ± 18.0 | | - | 82.7 ± 17.5 | | - | 82.7 ± 17.5 | | - |
| MSP score (points) | 12.1 ± 1.0 | 13.6 ± 1.5 | <0.001 | 12.1 ± 1.0 | 13.3 ± 1.7 | <0.001 | 12.1 ± 1.0 | 13.3 ± 1.7 | <0.001 |
| Grip strength (kg) | 23.2 ± 6.6 | 22.3 ± 6.7 | 0.002 | 23.4 ± 6.5 | 22.4 ± 6.7 | 0.001 | 23.5 ± 6.6 | 22.5 ± 6.8 | 0.002 |
| Male (kg) | 33.6 ± 5.7 | 32.7 ± 6.2 | 0.593 | 33.2 ± 5.6 | 31.8 ± 7.4 | 0.344 | 33.2 ± 5.6 | 31.8 ± 7.4 | 0.344 |
| Female (kg) | 20.6 ± 3.4 | 19.6 ± 3.3 | <0.001 | 20.6 ± 3.2 | 19.7 ± 3.2 | <0.001 | 20.5 ± 3.2 | 19.7 ± 3.3 | 0.001 |
| Sit-to-stand test (n/30 s) | 16.0 ± 5.7 | 17.3 ± 5.9 | 0.040 | 15.8 ± 5.4 | 17.0 ± 5.7 | 0.056 | 15.7 ± 5.5 | 17.1 ± 5.8 | 0.026 |
| Anteflexion on sitting (cm) | 43.7 ± 11.4 | 45.6 ± 10.2 | 0.349 | 41.7 ± 12.7 | 44.1 ± 10.9 | 0.221 | 41.7 ± 12.9 | 44.4 ± 10.8 | 0.141 |

Data are presented as mean ± standard deviation or number.

[a] Results were similar even after excluding individuals with low attendance (≤ one-third of the scheduled sessions).

Abbreviations: MSP, monowasure soudan proguramu (forgetfulness consultation program)—a computerized test battery for Alzheimer's disease screening (commercially available from Nihon Kohden Corporation)

Table S6. Participant characteristics and physical function test results in the comparison of pre- and post-intervention assessments (Yurihama Town)

| | Analysis of unique individuals | | | Analysis of total observations | | | Analysis excluding individuals with less participation [a] | | |
|---|---|---|---|---|---|---|---|---|---|
| | Pre | Post | P value | Pre | Post | P value | Pre | Post | P value |
| Number of individuals (n) | 27 | | - | 29 | | - | 24 | | - |
| Age (year) | 80.3 ± 5.7 | | - | 80.3 ± 5.6 | | - | 80.2 ± 6.1 | | - |
| Sex (male:female) (n) | 7:20 | | - | 7:22 | | - | 7:17 | | - |
| Class participation (Total 24 sessions) | | | | | | | | | |
| Frequency (times) | 17.5 ± 5.7 | | - | 16.5 ± 6.5 | | - | 18.8 ± 4.5 | | - |
| Percentage (%) | 72.8 ± 23.6 | | - | 68.8 ± 27.3 | | - | 78.3 ± 18.6 | | - |
| MSP score (points) | 11.8 ± 1.2 | 12.9 ± 2.3 | 0.046 | 11.9 ± 1.2 | 13.0 ± 2.2 | 0.031 | 11.6 ± 1.2 | 12.7 ± 2.4 | 0.071 |
| Grip strength (kg) | 21.3 ± 4.3 | 22.5 ± 5.1 | 0.052 | 21.3 ± 4.3 | 22.5 ± 5.1 | 0.052 | 21.4 ± 4.4 | 22.6 ± 5.1 | 0.054 |
| Male (kg) | 25.6 ± 2.2 | 28.3 ± 3.7 | 0.039 | 25.6 ± 2.2 | 28.3 ± 3.7 | 0.039 | 25.6 ± 2.2 | 28.3 ± 3.7 | 0.039 |
| Female (kg) | 20.0 ± 3.9 | 20.8 ± 4.0 | 0.292 | 20.0 ± 3.9 | 20.8 ± 4.0 | 0.292 | 20.1 ± 4.0 | 20.8 ± 4.2 | 0.301 |
| Gait speed (m/sec) | 1.6 ± 0.6 | 1.5 ± 0.5 | 0.812 | 1.6 ± 0.6 | 1.5 ± 0.5 | 0.812 | 1.6 ± 0.6 | 1.6 ± 0.5 | 0.824 |
| One-leg standing time (s) | 20.8 ± 18.3 | 21.3 ± 20.8 | 0.751 | 20.8 ± 18.3 | 21.3 ± 20.8 | 0.751 | 21.2 ± 18.6 | 21.3 ± 21.3 | 0.615 |

Data are presented as mean ± standard deviation or number.

[a] Results were similar for both analyses using the unique individual data (only one dataset per individual) and total observations data (using multiple data points even for a single individual).

Abbreviations: MSP, monowasure soudan proguramu (forgetfulness consultation program)—a computerized test battery for Alzheimer's disease screening (commercially available from Nihon Kohden Corporation)

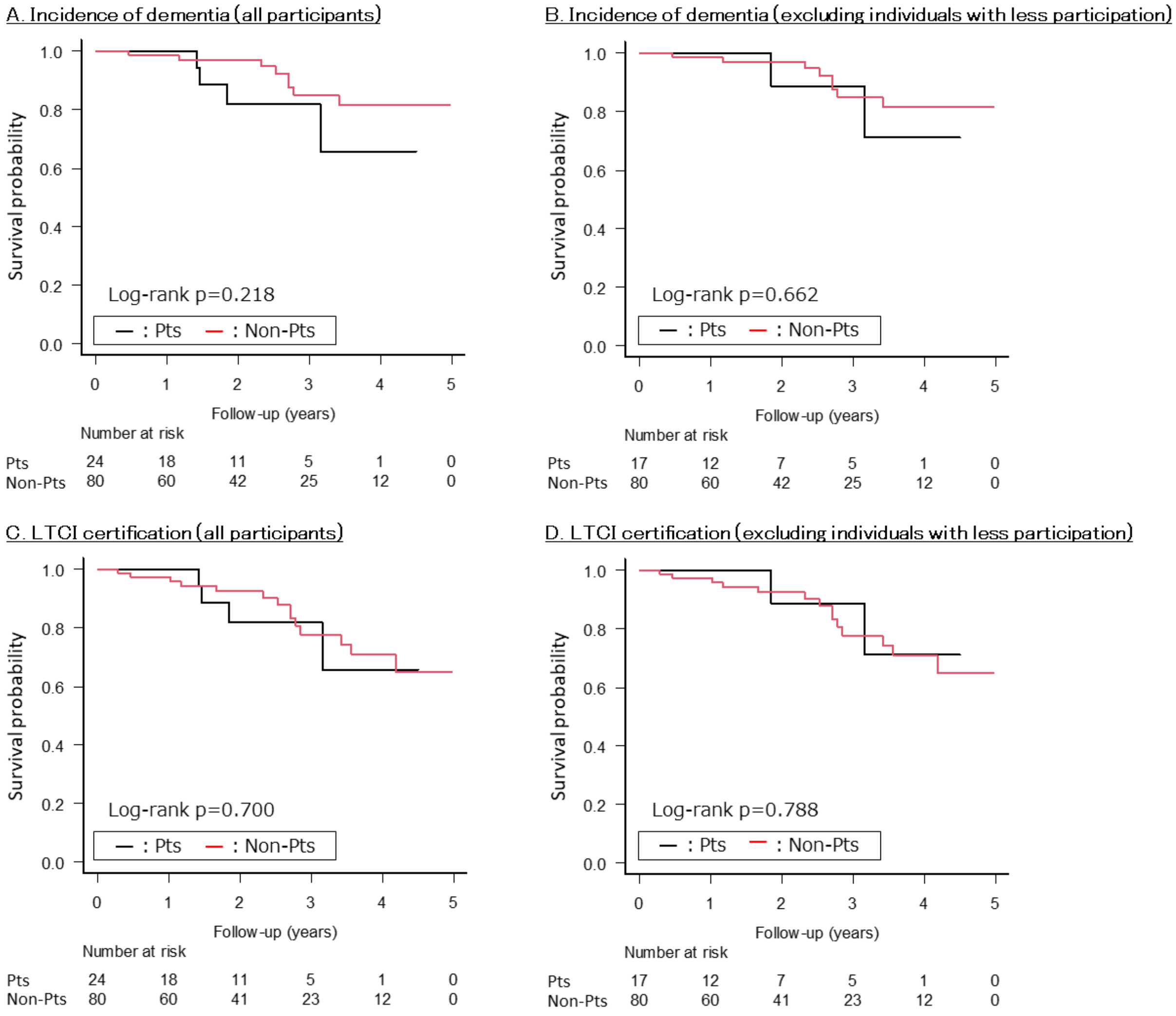


Figure S1. Kaplan–Meier curves for dementia incidence and long-term care insurance certification among participants and non-participants in the dementia prevention classes (Misasa Town).

Results for A: incidence of dementia (all participants), B: incidence of dementia (excluding individuals with less participation), C: LTCI certification (all participants), and D: LTCI certification (excluding individuals with less participation) are shown. Individuals with less participation were defined as those with low attendance (defined as attendance of one-third or less of the scheduled sessions) or discontinued participation (defined as absence for three or more consecutive sessions with no subsequent return).

Abbreviations: Pts, Participants; Non-Pts, Non-Participants; LTCI, long-term care insurance

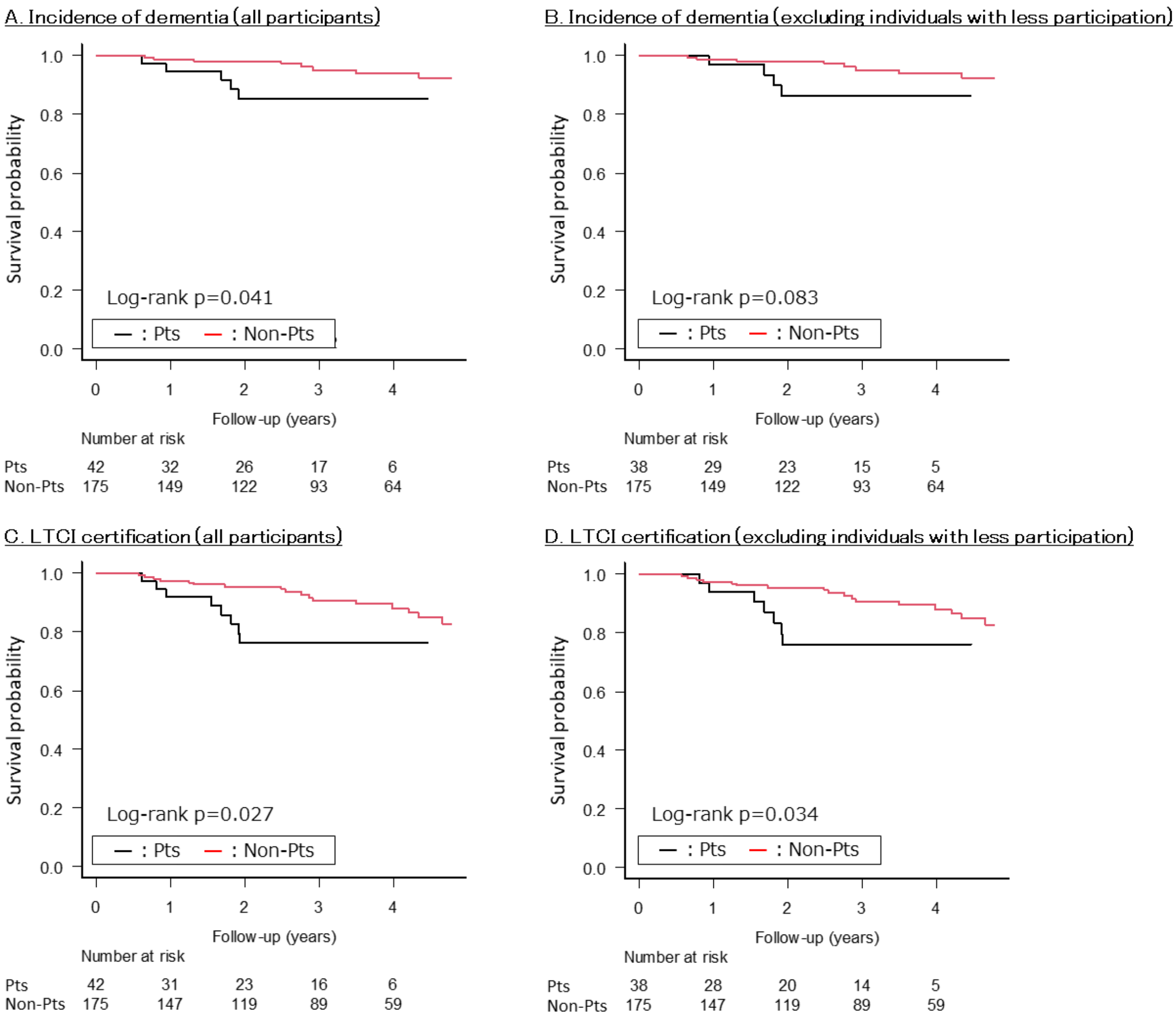


Figure S2. Kaplan–Meier curves for dementia incidence and long-term care insurance certification among participants and non-participants in the dementia prevention classes (Hokuei Town).

One participant in the class was excluded from the analysis due to missing data.

Results for A: incidence of dementia (all participants), B: incidence of dementia (excluding individuals with less participation), C: LTCI certification (all participants), and D: LTCI certification (excluding individuals with less participation) are shown. Individuals with less participation were defined as those with low attendance (defined as attendance of one-third or less of the scheduled sessions) or unknown attendance frequency.

Abbreviations: Pts, Participants; Non-Pts, Non-Participants; LTCI, long-term care insurance

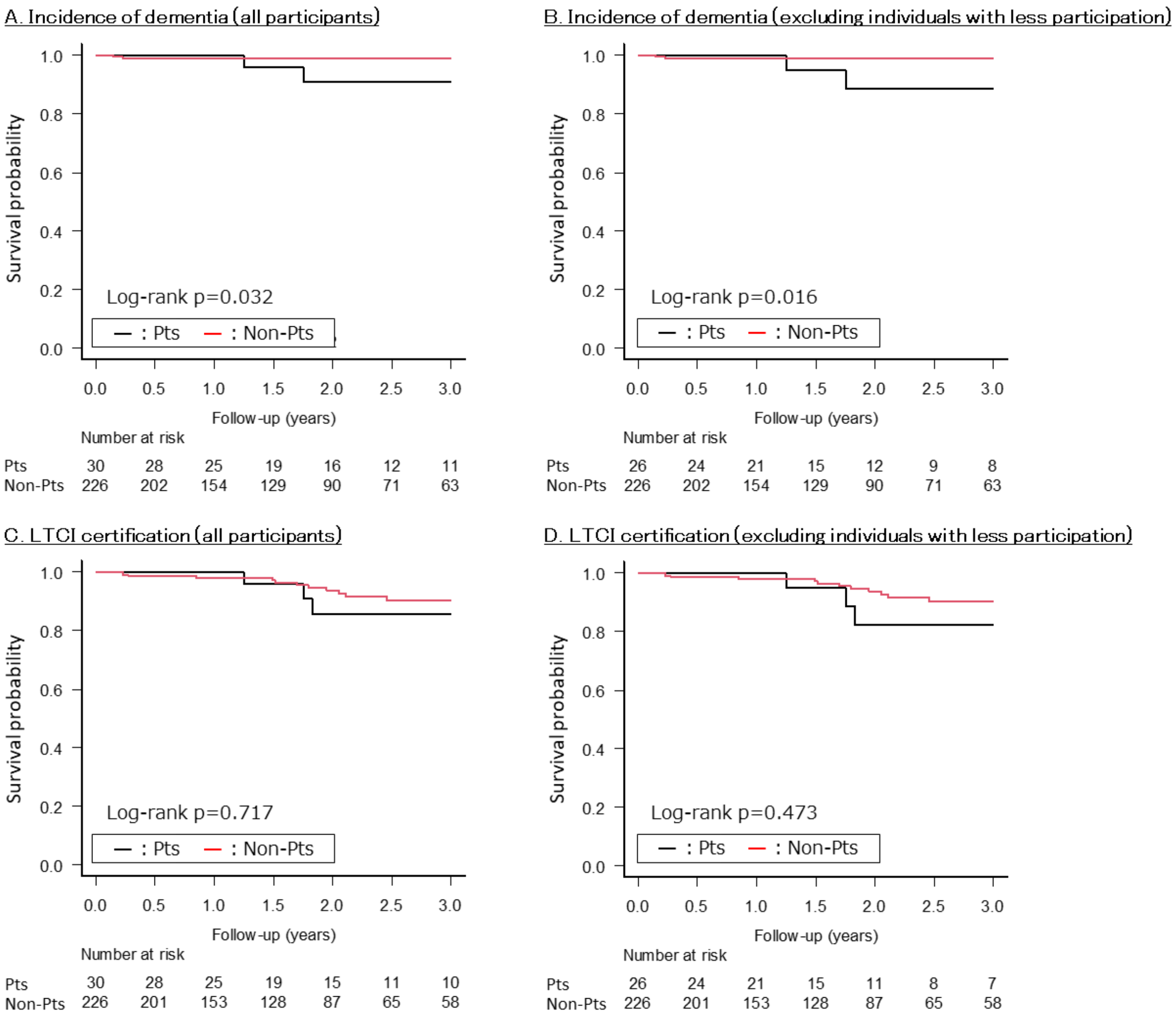


Figure S3. Kaplan–Meier curves for dementia incidence and long-term care insurance certification among participants and non-participants in the dementia prevention classes (Yurihama Town).

Results for A: incidence of dementia (all participants), B: incidence of dementia (excluding individuals with less participation), C: LTCI certification (all participants), and D: LTCI certification (excluding individuals with less participation) are shown. Individuals with less participation were defined as those with low attendance (attendance of one-third or less of the scheduled sessions).

Abbreviations: Pts, Participants; Non-Pts, Non-Participants; LTCI, long-term care insurance
